# Insurance coverage, healthcare use, and outpatient payment among adults with disability in Kenya before the SHA transition: a secondary analysis of KDHS 2022

**DOI:** 10.64898/2026.09.20.26363504

**Authors:** Nichodemus Werre Amollo, Japheth Ogol

## Abstract

**Background:** Adults with disability tend to need more healthcare and to have less capacity to pay for it. Kenya replaced the National Hospital Insurance Fund (NHIF) with the Social Health Authority (SHA) in October 2024, so KDHS 2022 is the last pre-transition national survey containing these measures. **We examined differences by disability status in health insurance coverage, healthcare use, and outpatient payment before the transition, including how insurance coverage varied across disability severity and what could be observed about insurer contributions at the point of care.**

**Methods:** Cross-sectional analysis of the KDHS 2022 person recode, covering adults aged 18 and above in the long-questionnaire subsample that carried the disability, insurance and service-use modules. Disability used the Washington Group Short Set (WG-SS). Survey-weighted quasi-Poisson models gave prevalence ratios under three adjustment sets with marginal standardised differences; severity categories were contrasted directly; amounts and payer sources were analysed among those who reported paying, with bounds where payer source is unrecorded.

**Results:** Of 40,197 adults, 6.7% met the WG threshold. Uninsurance was higher among them unadjusted (PR 1.10) and after adjustment for sex, age group and residence (PR 1.10, 95% CI 1.06 to 1.13; +6.7 percentage points), and was attenuated after adding wealth and education (PR 1.01, 95% CI 0.98 to 1.04; 0.6 percentage points, 95% CI -1.3 to 2.5). Outpatient use and hospitalisation were higher in every specification (fully adjusted PR 1.63, 14.0 percentage points, 95% CI 11.7 to 16.3; and PR 2.15, 7.7 percentage points, 95% CI 5.9 to 9.6; both p < 0.001). Among outpatient users with disability, 90.5% reported paying, insured or not. Among respondents who reported paying, insurance was associated with a lower probability that any part of the payment was made in cash (PR 0.89, p < 0.001), while among those who did pay cash the conditional cash amount was higher. Among insured outpatient users with disability who reported paying and had a recorded payer split, 15.1% had a recorded insurer contribution. Because payer information was not collected when no payment was reported, the corresponding proportion among all insured outpatient contacts was bounded between 11.9% and 32.9%.

**Conclusions:** Adults with disability had higher crude uninsurance, but this difference was attenuated to the null after adjustment for wealth and education. They also had substantially higher outpatient use and hospitalisation, which persisted under every adjustment set. Most outpatient users reported making a payment at their last visit regardless of insurance status, while the insurer contribution could only be partially observed because payer information was not collected when no payment was reported. These estimates provide a pre-transition benchmark, against which SHA-era measurement should assess payment at the point of care alongside insurance coverage.

## 1. Background

Disability is an under-measured dimension of health-system inequity. The World Health Organization estimates that about 1.3 billion people, roughly 16% of the world’s population, live with significant disability, and that barriers to care are steepest in low- and middle-income countries [1]. The financing implication is direct: adults with disability tend to need more care, while functional difficulty can constrain schooling, earnings and the ability to pay a contributory premium.

Kenyan evidence has established that insurance coverage is socially patterned and pro-rich. Analyses of earlier KDHS rounds documented large differences by wealth, education and employment [2,3], and a 36-country analysis found the same pattern wherever contributory mechanisms dominate [4]. Two analyses of KDHS 2022 itself are in print. One reports coverage levels and determinants among respondents aged 15 to 54 without examining disability; it analyses the individual and men’s recodes, where the standard insurance variables are empty in this round, and does not name the variable used, so its estimates are not directly comparable with ours [5]. The other uses coarsened exact matching to separate gross healthcare expenditure from net out-of-pocket payment after reimbursement [6]. Qualitative work with women with disabilities living in poverty in Kenya has shown that pro-poor financing policies can reach the enrolment margin and still leave people paying [7]. What is missing is a national, adult, disability-stratified account that places coverage, care-seeking and payment in one sample.

The policy context has since changed. The Social Health Insurance Act 2023 repealed the NHIF Act and established the Social Health Authority together with three funds: the Primary Healthcare Fund, the Social Health Insurance Fund and the Emergency, Chronic and Critical Illness Fund [8]. The Act does not create a disability-specific fund or a standing premium exemption for disability. It defines a person with disability who is wholly dependent on and living with a contributor as a beneficiary, includes persons with disability among vulnerable persons, and provides that contributions for households identified as unable to pay, through a means-testing instrument, are met from funds appropriated by Parliament [8,9]. Eligibility for that support therefore runs through household means testing and administrative identification, not through a functional-difficulty score. The separate legal architecture for disability certification, formerly the Persons with Disabilities Act 2003, was replaced by the Persons with Disabilities Act 2025, in force from 27 May 2025 [10,11]. KDHS 2022 records neither certification status nor programme enrolment, so this study cannot speak to who would qualify for public support under either instrument.

KDHS 2022 carries the Washington Group Short Set (WG-SS) in the person recode file, allowing functional difficulty to be examined alongside insurance coverage, healthcare use and payment [12,13]. **Against this background, there is a need for a pre-transition national description of whether adults with disability differ from other adults in insurance coverage and healthcare use, and whether insurance coverage translates into reduced payment at the point of care.**

**We therefore examined three related questions among Kenyan adults before the SHA transition:** (i) how insurance coverage, outpatient use and hospitalisation differed between adults meeting and not meeting the WG-SS disability threshold, both unadjusted and under prespecified adjustment sets; (ii) whether uninsurance varied across WG-SS severity categories; and (iii) among outpatient users, what was paid at the last visit, how much was paid, and what could be observed about the sources of payment, including insurer contributions. **Because the study is cross-sectional, these analyses describe associations and do not estimate causal effects of disability or insurance.**

### 1.1 Conceptual Framework

The conceptual framework in Figure 1 summarises the analytical structure of the study. At the population level, we examine the relationships of disability and functional difficulty with health insurance coverage and healthcare use, while considering key sociodemographic and socioeconomic characteristics. Among adults who reported recent outpatient use, we then examine whether a payment was reported, the amount paid and the payer sources recorded by the survey. This second stage is conditional on healthcare use and on the survey’s payment questions. Because payer information is not collected when respondents report no payment, insurer contribution across all insured outpatient contacts is only partially identified and is therefore bounded rather than estimated as a single proportion.

**Figure 1.**
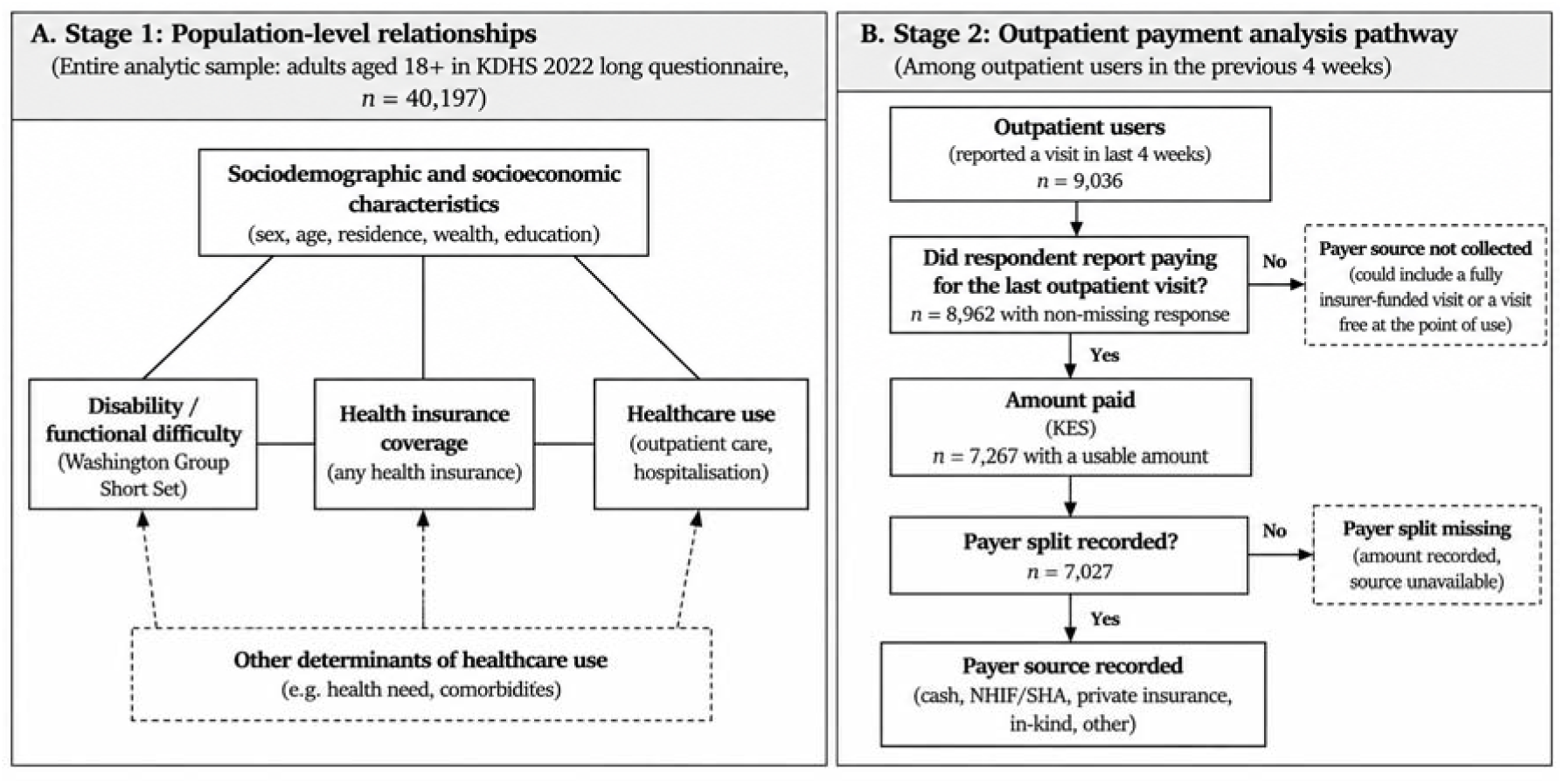
Conceptual framework for the descriptive analysis of disability, health insurance coverage, healthcare use, and outpatient payment among Kenyan adults. Note: The framework summarises the principal relationships examined in this cross-sectional analysis rather than specifying a causal pathway. Disability, sociodemographic and socioeconomic characteristics, and healthcare need may be interrelated and associated with insurance coverage and healthcare use. Outpatient payment is assessed only among respondents who reported healthcare use. Payer information is not collected when no payment is reported, so a fully insurer-funded visit cannot be distinguished from a visit that was free at the point of use. The temporal ordering of these factors cannot be established from KDHS 2022.

## 2. Methods

### 2.1 Study design and data source

This study is reported in accordance with the STROBE statement for cross-sectional studies.

KDHS 2022 used a stratified two-stage cluster design: enumeration areas were selected in the first stage and households within them in the second, with fieldwork conducted between February and July 2022 [12]. Within each cluster the sampled households were divided between two instruments: one in every two households received the full household questionnaire and the remainder a short version. The disability, insurance, service-use and expenditure modules analysed here were administered only in the full-questionnaire households, so the analytic base is that half-sample. All analyses use the person recode (PR) file, which holds one record per household member.

The household questionnaire is answered by a household respondent on behalf of household members. The disability, insurance, utilisation and payment items used here are therefore household-reported and, for most adults in the file, proxy-reported rather than self-reported. The PR file does not identify which member supplied the information for which record.

### 2.2 Study population

The analytic population was adult usual residents aged 18 years and above in long-questionnaire households with complete six-domain WG-SS data. The age threshold of 18 was set a priori to match the adult policy population for premium contribution, rather than the 15-year threshold used in the KDHS report tables. Participant flow from the raw file to each outcome-specific denominator is given in Table S1, and missing values by variable in Table S2. Analyses are complete-case within each outcome and model. Because the disability, insurance, utilisation and payment modules were administered only in long-questionnaire households, all estimates are restricted to that subsample, and the analysis does not attempt to reconstruct population totals from the full KDHS sample. The supplied person weights were retained for estimation and inference was based on the parent survey design. Because the recode documentation does not establish a separate calibration adjustment for the questionnaire split, the results should be read as weighted estimates for the long-questionnaire population rather than as independently recalibrated national totals. Table S7 compares the two halves on the five characteristics recorded in both, as a check on how far the restriction moves the composition of the sample.

### 2.3 Variables

#### 2.3.1 Disability

The WG-SS comprises six functional-difficulty items, captured in KDHS 2022 as seeing (hdis2), hearing (hdis4), communication in the respondent’s usual language (hdis5), remembering or concentrating (hdis6), walking or climbing steps (hdis7) and self-care (hdis8) [13]. The adjacent variables hdis1 and hdis3, which in the standard recode carry use of glasses and of hearing aids, are empty in this dataset and were not used; the seeing and hearing items were therefore analysed as recorded, without information on whether the respondent was assessed with usual assistive devices. The WG threshold was defined as “a lot of difficulty” or “cannot do at all” in at least one domain.

Severity was taken from hdis9, the highest degree of difficulty reported across the six domains, and labelled no, mild, moderate and severe functional difficulty. This is a maximum, not a scale: one severe domain and substantial difficulty in several domains are collapsed differently by it. The count of domains at or above threshold is therefore analysed as a prespecified alternative specification.

#### 2.3.2 Insurance, service use and payment

Insurance coverage was taken from sh27, an individual-level item recorded for each household member within the household questionnaire, and NHIF coverage from sh28a. Uninsurance was defined as a “no” response; the 286 adults answering “don’t know” were treated as missing in the main analysis and grouped with the uninsured in a prespecified sensitivity analysis. Outpatient use in the previous four weeks came from sh31. Hospitalisation came from sh29, which records at least one overnight stay in a health facility in the previous 12 months; the number of admissions is not analysed.

Payment was measured at three levels, on three nested populations that are kept distinct throughout because they have different denominators:

• **Outpatient users.** Whether any money was paid at the last outpatient visit came from sh32. This is an incidence measure and says nothing about magnitude.

• **Payers.** The total cost of that visit came from sh304, asked only of respondents who reported paying.

• **Payers with a payer split.** The amounts met in cash, by NHIF, by private insurance, in kind and by other means came from sh305a to sh305e.

Because the amount and payer items are skipped for anyone who reported no payment, the survey records nothing about the payer at a contact where no money changed hands, including one an insurer may have met in full. Estimates from the second and third populations therefore describe payments that were made and are not generalised to all contacts. This missing-by-design structure means the observed payer data cannot identify the proportion of all insured outpatient contacts at which an insurer contributed: a visit an insurer paid in full is indistinguishable from a visit that was free at the point of care. We therefore treat insurer contribution as a partially identified quantity and report bounds rather than a single point estimate (Table S4). Both endpoints of that bound are population proportions and are estimated with the survey weights on the parent design, with design-based limits reported for each endpoint separately.

sh305a is the amount reported as met in cash at the visit. The questionnaire does not record reimbursement received afterwards, so this is described as the cash paid at the point of care rather than as a net payment after reimbursement. Values coded 999998 and above are non-response codes and were set to missing. KDHS does not collect household consumption or income, so catastrophic and impoverishing health expenditure cannot be constructed from this file, and no such measure is reported.

The payer breakdown is itself incomplete: 187 respondents with a usable total have no payer split at all, and the split is recorded asymmetrically for a further handful, with a cash amount present and the NHIF field missing. Those records are excluded from the payer columns and retained in the amount columns, which is why the two carry different denominators.

Three internal inconsistencies in the amount items were counted rather than silently carried: 186 records report a payment with a total of zero, 5 have payer components summing above the reported total, and 4 have a cash amount above the reported total. A prespecified sensitivity analysis excludes all of them.

#### 2.3.3 Covariates and adjustment sets

Three specifications are reported for each whole-sample contrast: unadjusted; adjusted for sex, age group (18-29, 30-44, 45-59, 60+) and place of residence (hv025); and additionally adjusted for wealth quintile (hv270) and educational attainment (hv106).

The specifications are named for what they adjust for rather than for a causal role. KDHS 2022 records no age at onset of functional difficulty, so the temporal ordering of education, wealth and disability cannot be established from these data. For difficulty acquired in later life, education is prior to the exposure and would confound it; for lifelong or early-onset difficulty, schooling and earning capacity are plausibly downstream of it, and adjusting for them would remove part of the association of interest. Current wealth may be both a cause and a consequence of functional limitation, and residence may change after onset. A difference between the second and third specification is therefore reported as attenuation, which is consistent with confounding, with mediation, with measurement error in the adjustment variables, or with a combination; it is not reported as a mediated effect.

County (hv024) was not entered in the models: the long-questionnaire subsample is not powered for 47-county fixed effects in the disability stratum, and no county-level estimate is reported.

### 2.4 Statistical analysis

All estimates used the complex design: PSU (hv021), stratum (hv022) and household-member weight (hv005/1,000,000), with nest = TRUE. One parent design was built for all adult usual residents, and every subgroup estimate was taken as a domain of it with subset(), so that variances retain the parent PSU and stratum structure, including PSUs contributing no observations to a domain. The analytic base contains 1,691 PSUs in 92 strata, giving 1,599 design degrees of freedom. Strata containing a single PSU were handled with the centring adjustment (options(survey.lonely.psu = "adjust")). Confidence intervals for proportions were logit-transformed; intervals for regression coefficients and contrasts use the survey degrees of freedom.

Prevalence ratios were estimated with survey-weighted quasi-Poisson models with a log link, appropriate where the outcome is common [14]. For each whole-sample outcome we report the prevalence ratio, the standardised prevalence under each exposure setting, and the marginal standardised difference in percentage points, computed by g-computation over the covariate distribution of the analytic sample. Delta-method limits for those differences are model-based, in that they treat the weighted covariate distribution as fixed; they are therefore checked against limits from 500 survey-design bootstrap replicates in which the model is refitted for each replicate. Results were materially unchanged between the two variance estimators (Table S6), whose construction is described in the supplementary material.

Severity was tested with a prespecified family of four contrasts, the three adjacent steps and the extreme contrast of severe against no difficulty, estimated on the linear predictor and exponentiated, together with a design-based global Wald test of the severity term. The contrasts were prespecified to describe the pattern across the ordered categories and were not treated as confirmatory hypothesis tests, so no multiplicity adjustment was applied. Overlapping confidence intervals were not used as a test, and no equivalence claim is made, since no equivalence margin was prespecified.

Amounts were summarised as survey-weighted medians and means with confidence intervals. The cash payment was modelled in two parts, because zero cash is one of the two ways insurance can appear in these data and is concentrated among insured respondents: part one is the probability of paying anything in cash, and part two is the amount among those who paid something, which is conditional on that. Modelling only part two would exclude the zero-cash records and remove the observations most likely to be consistent with an insurance contribution. The gross amount model is likewise conditional on a payment having been made. Both use a survey-weighted linear model on the log amount, given the strong right skew, and are reported as ratios of geometric means.

Two analyses condition on having used outpatient care or on having paid. Because disability, insurance, wealth and need all influence care-seeking, these are descriptive associations within the selected group and are not interpretable as effects of insurance; this is stated wherever they are reported.

Prespecified sensitivity analyses covered the coding of “don’t know” insurance responses, the multi-domain alternative to the severity summary, exclusion of the internally inconsistent amount records, and a comparison of adults included in and excluded from the fully adjusted model. Analyses followed DHS guidance on weighting and design-based estimation [15] and used R 4.5.1 with survey 4.4.2 [16]; full package versions and session information are deposited with the analysis code.

### 2.5 Ethical considerations

This study used de-identified secondary data from KDHS 2022, obtained from The DHS Program under an approved data request. The original survey was approved by the Kenya Medical Research Institute Scientific and Ethics Review Unit and the ICF Institutional Review Board. No new participant contact occurred and no additional ethical approval was required.

## 3. Results

### 3.1 Participant flow and sample characteristics

Of 156,571 records in the PR file, 77,909 were adult usual residents with complete design variables, of whom 37,625 were in short-questionnaire households where the modules were not administered. The analytic base was 40,197 adults in 19,695 long-questionnaire households with complete WG-SS data (Table S1). Long-questionnaire and short-questionnaire adults were closely similar on the five characteristics recorded in both halves, the largest weighted difference being 0.9 percentage points (Table S7). Item missingness within the analytic base was below 1% for every outcome and covariate (Table S2); adults excluded from the fully adjusted insurance model (393, 1.0%) were similar in age to those included (mean 39.7 versus 38.9 years) but less often rural (50% versus 65%).

Any functional difficulty was reported by 25.6% (95% CI 24.9% to 26.4%) of adults, and 6.7% (95% CI 6.3% to 7.0%) met the WG threshold. Adults at the threshold were much older than adults below it (mean age 57.6 (56.6, 58.5) versus 37.1 (36.8, 37.4) years), more often rural, and concentrated in the poorer quintiles (Table 1).

**Table 1.** Weighted sociodemographic characteristics of adults aged 18 years and above by Washington Group Short Set disability status, KDHS 2022.

| Characteristic | n | Overall<br>Estimate (95%<br>CI) | n | No WG disability<br>Estimate (95%<br>CI) | n | WG disability threshold<br>Estimate (95%<br>CI) |
| --- | --- | --- | --- | --- | --- | --- |
| Sample size, n | 40,197 |  | 37,364 |  | 2,833 |  |
| Mean age, years (95% CI) | 40,197 | 38.5 (38.2, 38.8) | 37,364 | 37.1 (36.8, 37.4) | 2,833 | 57.6 (56.6, 58.5) |
| Sex |  |  |  |  |  |  |
| Women | 21,386 | 52.8 (52.2, 53.4) | 19,695 | 52.2 (51.6, 52.8) | 1,691 | 60.6 (58.5, 62.6) |
| Age group |  |  |  |  |  |  |
| 18-29 | 14,756 | 37.4 (36.5, 38.2) | 14,399 | 39.1 (38.2, 40.0) | 357 | 12.9 (11.5, 14.4) |
| 30-44 | 12,583 | 31.9 (31.1, 32.7) | 12,149 | 33.0 (32.2, 33.9) | 434 | 15.7 (13.9, 17.6) |
| 45-59 | 7,159 | 17.6 (17.0, 18.1) | 6,584 | 17.3 (16.8, 17.9) | 575 | 20.5 (18.7, 22.5) |
| 60+ | 5,699 | 13.2 (12.7, 13.7) | 4,232 | 10.5 (10.0, 11.0) | 1,467 | 50.9 (48.6, 53.2) |
| Place of residence |  |  |  |  |  |  |
| Rural | 25,912 | 62.6 (61.3, 63.9) | 23,731 | 61.2 (59.9, 62.6) | 2,181 | 81.9 (79.2, 84.2) |
| Wealth quintile |  |  |  |  |  |  |
| Poorest | 9,261 | 16.4 (15.5, 17.3) | 8,348 | 15.6 (14.7, 16.5) | 913 | 27.6 (25.2, 30.1) |
| Poorer | 7,361 | 18.4 (17.6, 19.4) | 6,712 | 18.0 (17.1, 18.9) | 649 | 25.4 (23.1, 27.8) |
| Middle | 8,194 | 19.7 (18.8, 20.7) | 7,557 | 19.5 (18.6, 20.4) | 637 | 23.2 (21.1, 25.5) |
| Richer | 8,781 | 22.6 (21.2, 23.9) | 8,349 | 23.1 (21.7, 24.5) | 432 | 14.8 (12.8, 17.1) |
| Richest | 6,600 | 22.9 (21.0, 24.8) | 6,398 | 23.9 (22.0, 25.9) | 202 | 9.0 (7.1, 11.5) |
| Educational attainment |  |  |  |  |  |  |
| No education | 6,433 | 9.4 (9.0, 9.9) | 5,369 | 7.9 (7.4, 8.3) | 1,064 | 31.2 (29.0, 33.4) |
| Primary | 14,541 | 36.8 (35.8, 37.8) | 13,360 | 36.1 (35.1, 37.2) | 1,181 | 46.0 (43.7, 48.3) |
| Secondary | 13,032 | 35.1 (34.4, 35.9) | 12,588 | 36.4 (35.6, 37.3) | 444 | 17.4 (15.5, 19.6) |
| Higher | 6,077 | 18.6 (17.4, 19.9) | 5,943 | 19.6 (18.3, 20.9) | 134 | 5.4 (4.3, 6.8) |

Source: Kenya DHS 2022. Percentages are weighted column percentages; n is the unweighted count in that category. Restricted to adult usual residents in the long-questionnaire subsample with complete WG-SS classification.

### 3.2 Insurance coverage across the severity gradient

Unadjusted coverage fell across severity categories, from 31.7 (30.3, 33.1) at no difficulty to 19.7 (13.9, 27.0) at severe difficulty, with NHIF tracking any insurance closely at every level (Table 2). The steps between adjacent severity categories were tested directly rather than inferred from the intervals in Table 2. Unadjusted, uninsurance at mild difficulty did not differ from uninsurance at no difficulty (PR 1.00, 95% CI 0.98 to 1.03; p = 0.722), the step from mild to moderate did (PR 1.09, 95% CI 1.05 to 1.13; p < 0.001), and the further step from moderate to severe did not (PR 1.07, 95% CI 0.98 to 1.17; p = 0.110). After adjustment for sex, age group and residence, both the mild step (PR 1.03, 95% CI 1.01 to 1.06; p = 0.018) and the mild-to-moderate step (PR 1.07, 95% CI 1.03 to 1.11; p < 0.001) reached significance. The design-based global test of severity was significant unadjusted (F = 13.29 on 3 and 1,596 df, p < 0.001) and after that adjustment (F = 13.70 on 3 and 1,591 df, p < 0.001), but not after wealth and education were added (F = 0.17 on 3 and 1,584 df, p = 0.91), where every contrast in the family was null (Table S3). The multi-domain alternative behaved the same way: relative to no domain at threshold, the fully adjusted uninsurance APR was 1.00, 95% CI 0.97 to 1.03 for one domain and 1.03, 95% CI 0.99 to 1.07 for two or more (global p = 0.38).

**Table 2.**
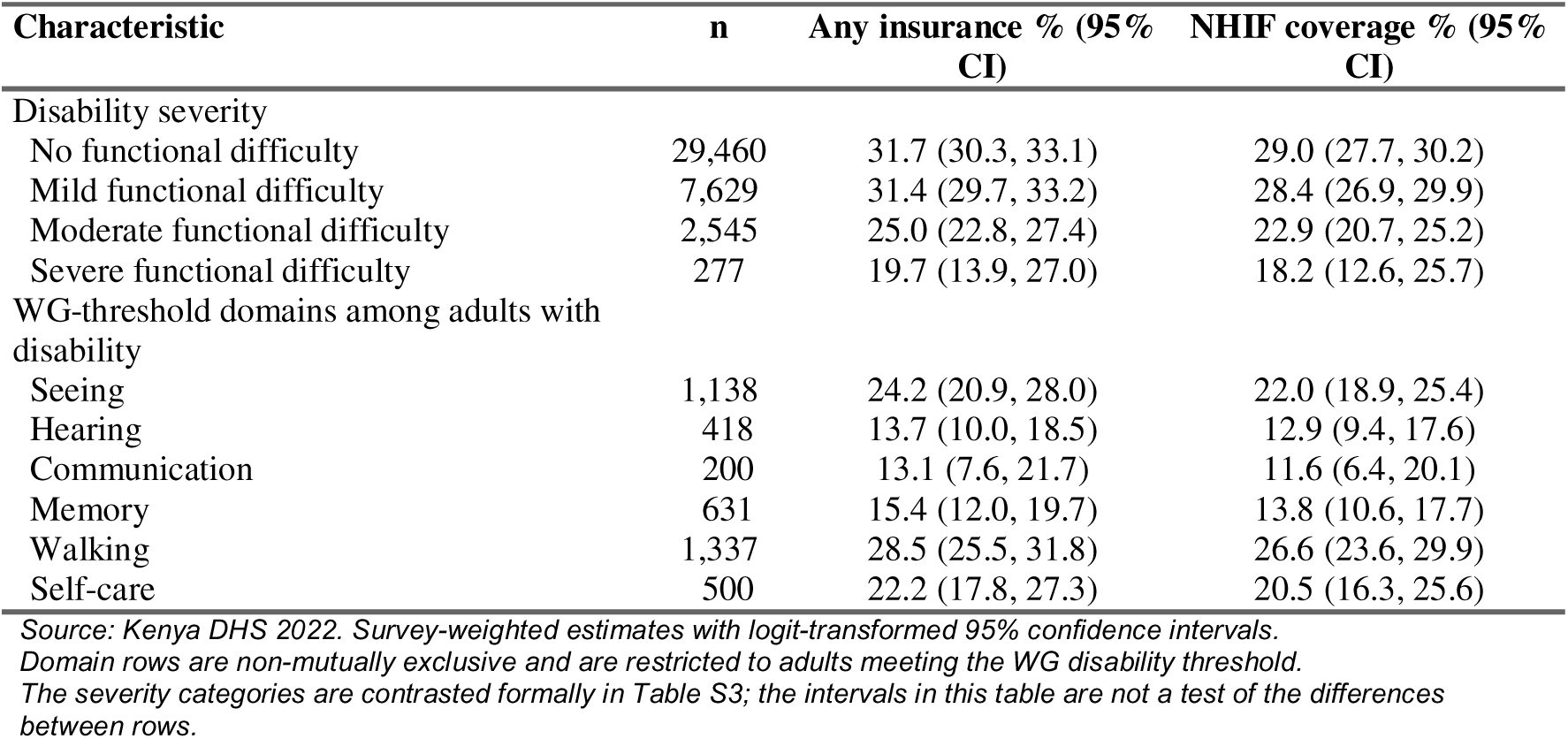
Weighted insurance coverage by Washington Group Short Set disability severity and functional domain, adults aged 18 years and above, KDHS 2022.

| Characteristic | n | Any insurance % (95% CI) | NHIF coverage % (95% CI) |
| --- | --- | --- | --- |
| Disability severity |  |  |  |
| No functional difficulty | 29,460 | 31.7 (30.3, 33.1) | 29.0 (27.7, 30.2) |
| Mild functional difficulty | 7,629 | 31.4 (29.7, 33.2) | 28.4 (26.9, 29.9) |
| Moderate functional difficulty | 2,545 | 25.0 (22.8, 27.4) | 22.9 (20.7, 25.2) |
| Severe functional difficulty | 277 | 19.7 (13.9, 27.0) | 18.2 (12.6, 25.7) |
| WG-threshold domains among adults with disability |  |  |  |
| Seeing | 1,138 | 24.2 (20.9, 28.0) | 22.0 (18.9, 25.4) |
| Hearing | 418 | 13.7 (10.0, 18.5) | 12.9 (9.4, 17.6) |
| Communication | 200 | 13.1 (7.6, 21.7) | 11.6 (6.4, 20.1) |
| Memory | 631 | 15.4 (12.0, 19.7) | 13.8 (10.6, 17.7) |
| Walking | 1,337 | 28.5 (25.5, 31.8) | 26.6 (23.6, 29.9) |
| Self-care | 500 | 22.2 (17.8, 27.3) | 20.5 (16.3, 25.6) |

Source: Kenya DHS 2022. Survey-weighted estimates with logit-transformed 95% confidence intervals.
Domain rows are non-mutually exclusive and are restricted to adults meeting the WG disability threshold.
The severity categories are contrasted formally in Table S3; the intervals in this table are not a test of the differences between rows.

NHIF coverage was within about three percentage points of any insurance at every severity level, so cover by private and community schemes was correspondingly small. Among adults at the WG threshold, coverage also varied by domain, from 13.1 (7.6, 21.7) for communication difficulty to 28.5 (25.5, 31.8) for walking difficulty; these domain estimates are descriptive, non-mutually exclusive and were not contrasted formally.

### 3.3 The disability contrast under three adjustment sets

Adults at the WG threshold were more likely to be uninsured than adults below it, unadjusted (PR 1.10, 95% CI 1.07 to 1.14) and after adjustment for sex, age group and residence (PR 1.10, 95% CI 1.06 to 1.13; standardised prevalence 75.0% versus 68.3%, a difference of 6.7 percentage points, 95% CI 4.4 to 9.0). After additionally adjusting for wealth quintile and educational attainment the association was attenuated to the null (PR 1.01, 95% CI 0.98 to 1.04; difference 0.6 percentage points, 95% CI -1.3 to 2.5). The attenuation appears only when those two variables enter: the estimate adjusted for sex, age group and residence is indistinguishable from the unadjusted one.

The two utilisation contrasts behaved differently. Outpatient use in the previous four weeks was higher among adults at the threshold in every specification (crude 1.92, 95% CI 1.80 to 2.04; adjusted for sex, age group and residence 1.56, 95% CI 1.46 to 1.67; with wealth and education added 1.63, 95% CI 1.52 to 1.74), as was hospitalisation in the previous 12 months (2.15, 95% CI 1.88 to 2.46 in the fullest specification, a difference of 7.7 percentage points, 95% CI 5.9 to 9.6). Table 3 reports the three outcomes together; replicate-weight limits for the standardised differences are in Table S6 and are close to the delta-method limits throughout.

**Table 3.** Prevalence ratios and standardised prevalence differences for adults meeting the Washington Group Short Set disability threshold compared with adults below it, whole adult sample, KDHS 2022.

| **Outcome and specification** | **Prevalence ratio (95% CI);**<br>**p** | **Standardised prevalence, %**<br>**(disability vs no disability)** | **Standardised difference, percentage points (95% CI)** |
| --- | --- | --- | --- |
| **Uninsured (n = 39,804)** |  |  |  |
| Crude | 1.10 (1.07, 1.14); p < 0.001 | 75.4 vs 68.3 | +7.1 (+4.7, +9.5) |
| Adjusted for sex, age group, residence | 1.10 (1.06, 1.13); p < 0.001 | 75.0 vs 68.3 | +6.7 (+4.4, +9.0) |
| Additionally adjusted for wealth, education | 1.01 (0.98, 1.04); p = 0.542 | 69.3 vs 68.7 | +0.6 (-1.3, +2.5) |
| **Outpatient use in previous 4 weeks (n = 39,953)** |  |  |  |
| Crude | 1.92 (1.80, 2.04); p < 0.001 | 42.1 vs 22.0 | +20.1 (+17.7, +22.5) |
| Adjusted for sex, age group, residence | 1.56 (1.46, 1.67); p < 0.001 | 34.8 vs 22.3 | +12.5 (+10.2, +14.7) |
| Additionally adjusted for wealth, education | 1.63 (1.52, 1.74); p < 0.001 | 36.2 vs 22.2 | +14.0 (+11.7, +16.3) |
| **Hospitalisation in previous 12 months (n = 40,028)** |  |  |  |
| Crude | 2.24 (1.98, 2.54); p < 0.001 | 15.0 vs 6.7 | +8.3 (+6.6, +10.0) |
| Adjusted for sex, age group, residence | 2.06 (1.80, 2.36); p < 0.001 | 13.9 vs 6.8 | +7.2 (+5.4, +9.0) |
| Additionally adjusted for wealth, education | 2.15 (1.88, 2.46); p < 0.001 | 14.5 vs 6.7 | +7.7 (+5.9, +9.6) |

*Source: Kenya DHS 2022. Survey-weighted quasi-Poisson models with a log link; confidence limits use the survey degrees of freedom.*
*Specifications are named for what they adjust for. KDHS records no age at disability onset, so whether education and wealth precede or follow functional difficulty cannot be established here; the change between the second and third row of each block is attenuation, not an estimated mediated effect.*

Figure 2 plots the same prevalence ratios with their confidence limits, one panel per outcome and one row per adjustment set. What it adds to Table 3 is the shape of the change as the adjustment set grows: in the uninsurance panel the interval moves onto the null once wealth and education enter, while in the two utilisation panels every interval stays well clear of it and the fully adjusted estimate sits no closer to the null than the crude one. The coverage contrast is an order of magnitude smaller than the hospitalisation contrast, so the panels are drawn on separate scales and should be read one at a time rather than compared against each other by eye.

**Figure 2.**
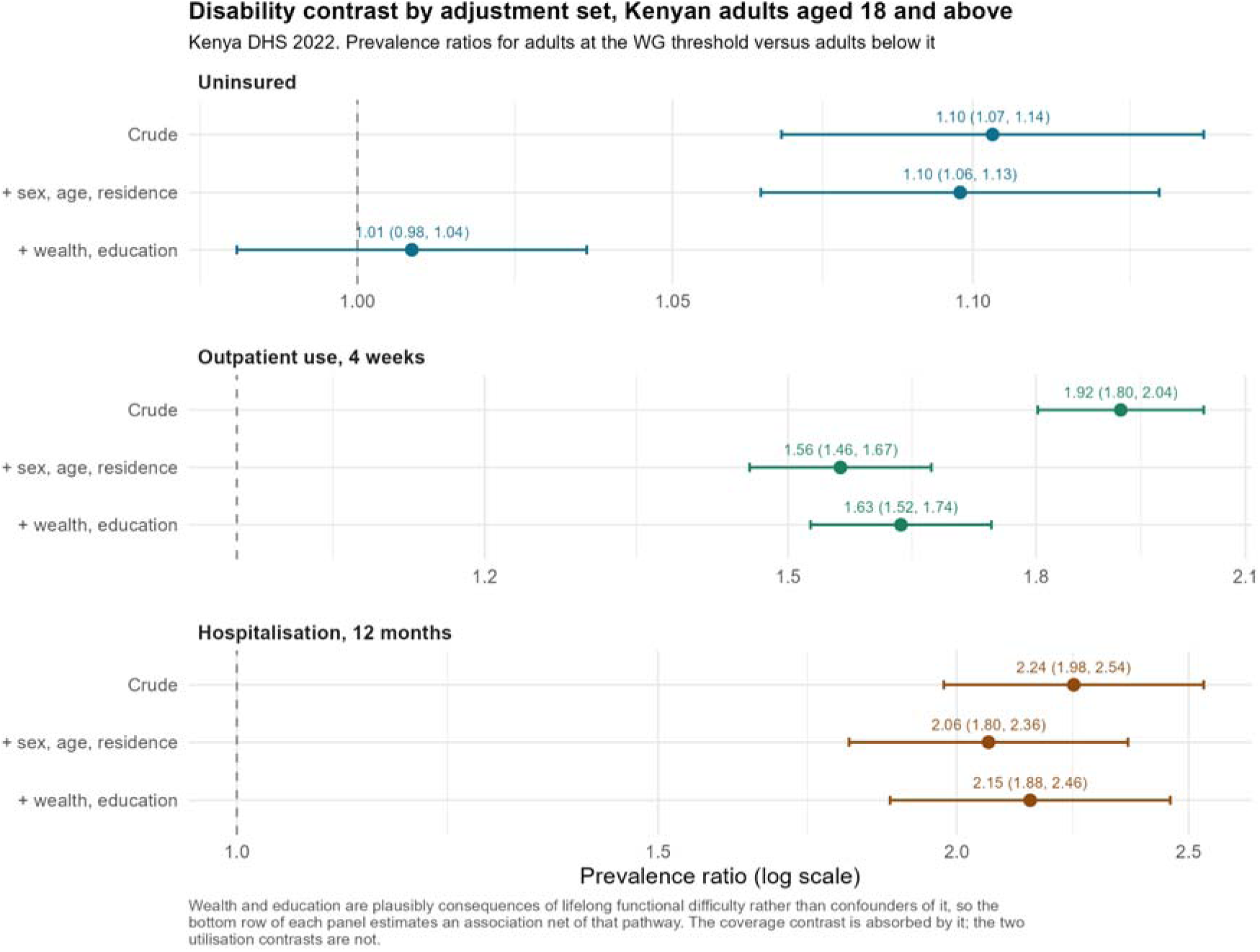
Prevalence ratios for adults at the Washington Group disability threshold versus adults below it, by adjustment set, KDHS 2022. Points show prevalence ratios and horizontal bars 95% confidence intervals on a log scale. Each panel is drawn on its own sc

**Figure 3.**
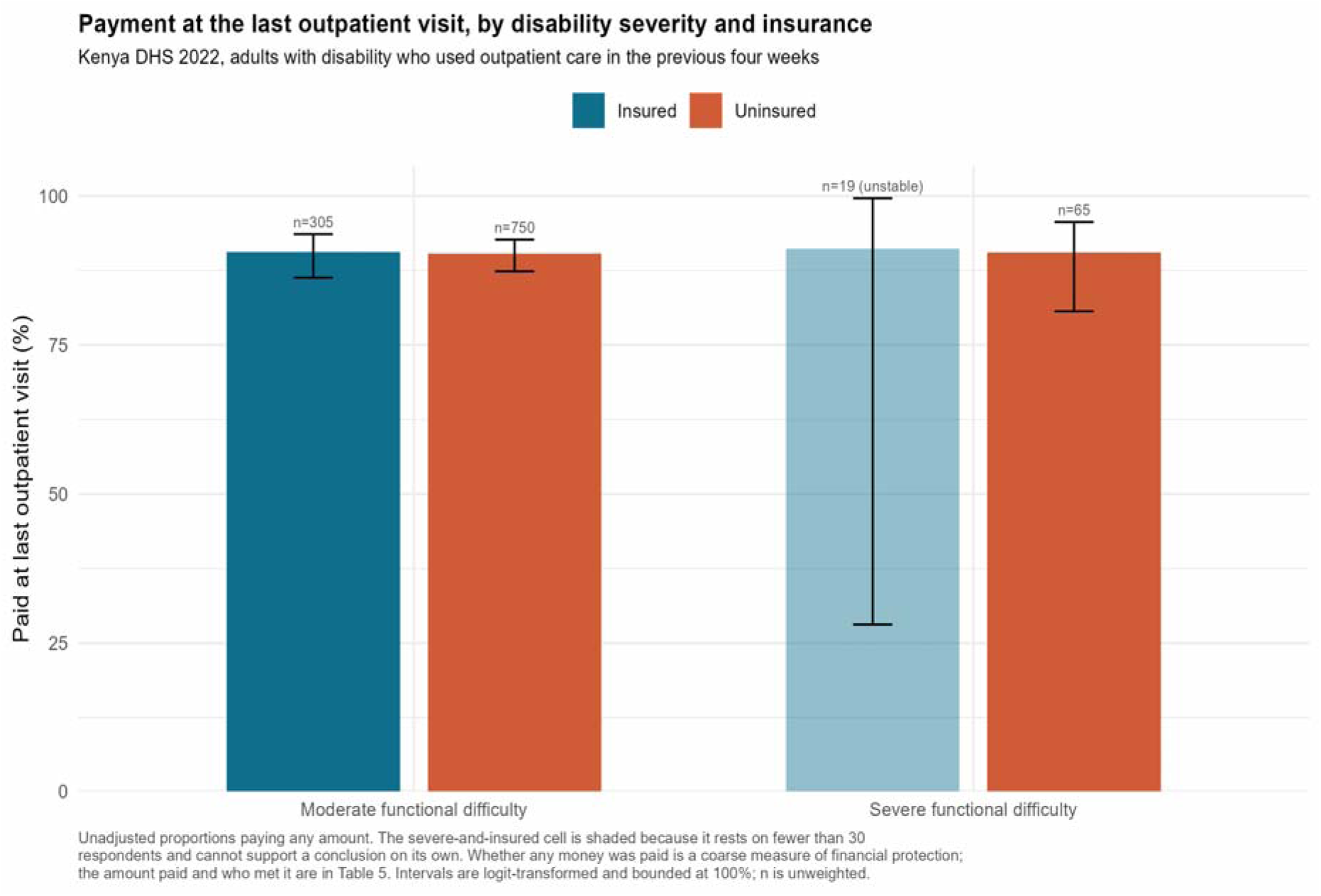
Payment at the last outpatient visit by disability severity and insurance status among adult outpatient users with disability in Kenya, KDHS 2022. Bars show survey-weighted proportions and error bars logit-transformed 95% confidence intervals; the shad

### 3.4 Service use and payment at outpatient contact

Outpatient use was 42.0% (95% CI 39.7% to 44.4%) among adults at the WG threshold and 22.0% (95% CI 21.3% to 22.7%) among adults below it; hospitalisation was 15.0% (95% CI 13.4% to 16.8%) and 6.7% (95% CI 6.3% to 7.1%) respectively (Table 4).

**Table 4.** Weighted service utilisation and payment at the last outpatient visit by disability status and insurance coverage, adults aged 18 years and above, KDHS 2022.

| Analysis group | n | Outpatient use<br>% (95% CI) | n | Hospitalisation<br>% (95% CI) | n | Paid at last<br>visit % (95%<br>CI) |
| --- | --- | --- | --- | --- | --- | --- |
| Disability status |  |  |  |  |  |  |
| No WG disability | 37,233 | 22.0 (21.3,<br>22.7) | 37,309 | 6.7 (6.3, 7.1) | 7,820 | 87.7 (86.6, 88.8) |
| WG disability threshold | 2,830 | 42.0 (39.7,<br>44.4) | 2,831 | 15.0 (13.4,<br>16.8) | 1,142 | 90.5 (88.2, 92.3) |
| Insurance status,<br>outpatient users with<br>disability |  |  |  |  |  |  |
| Insured |  |  |  |  | 324 | 90.6 (86.6, 93.5) |
| Uninsured |  |  |  |  | 815 | 90.4 (87.5, 92.6) |

Source: Kenya DHS 2022. Survey-weighted estimates; $n$ is the unweighted denominator for the adjacent column.
Utilisation is estimated among all adults in the row group; payment only among those who used outpatient care in the previous four weeks.
Payment records whether any money was paid, not how much. Amounts and payers are in Table 5.

Among adults who used outpatient care, payment of some amount was close to universal: 90.5% (95% CI 88.2% to 92.3%) of those with disability and 87.7% (95% CI 86.6% to 88.8%) of those without. Within the disability group, payment incidence was 90.6% (95% CI 86.6% to 93.5%) among insured users and 90.4% (95% CI 87.5% to 92.6%) among uninsured users (Table 4). In the whole-sample model among outpatient users, holding any insurance was not associated with a lower probability of reporting a payment (APR 0.98 (95% CI 0.96 to 1.00); p = 0.111). Payment was reported by 88% of users overall.

Figure 2 divides that comparison along a dimension Table 4 does not: among outpatient users with disability it sets insured against uninsured separately at moderate and at severe functional difficulty. The four proportions fall within a few percentage points of one another, so neither insurance nor severity marks out who paid something at the last visit. The insured severe cell rests on 19 respondents and its interval runs from 28% to 100%; it is shaded for that reason and carries nothing on its own.

### 3.5 What was paid, and who paid it

Among the 7,228 adults who reported paying and gave a usable total, the median cost of the visit was 1,500 KSh for insured adults with disability and 500 KSh for uninsured adults with disability (Table 5). Adjusted for sex, age group, residence, wealth and education, and conditional on a payment having been made, adults with disability paid 1.37 (95% CI 1.19 to 1.57) times as much as adults without disability (n = 7,032) and insured adults 1.40 (95% CI 1.25 to 1.58) times as much as uninsured adults. Excluding the internally inconsistent amount records left these estimates unchanged (1.36, 95% CI 1.19 to 1.56; and 1.40, 95% CI 1.25 to 1.58).

**Table 5.** Amount paid at the last outpatient visit and the source of that payment, among adults who used outpatient care in the previous four weeks and reported paying, KDHS 2022.

| Group | n with amount | Median total cost, KSh (95% CI) | Paid nothing in cash, % (95% CI) | Mean cash paid, KSh (95% CI) | n with payer split | Insurer met any part, % (95% CI) |
| --- | --- | --- | --- | --- | --- | --- |
| No WG disability, uninsured | 4,200 | 500 (500, 600) | 0.7 (0.4, 1.2) | 1,393 (1,139, 1,646) | 4,113 | 0.1 (0.0, 0.3) |
| No WG disability, insured | 2,082 | 750 (650, 900) | 14.7 (12.7, 17.0) | 1,657 (1,385, 1,929) | 1,986 | 18.0 (15.7, 20.4) |
| WG disability, uninsured | 691 | 500 (500, 680) | 1.2 (0.6, 2.6) | 2,335 (1,902, 2,767) | 683 | 0 (not estimable) |
| WG disability, insured | 255 | 1,500 (1,000, 2,000) | 11.5 (7.3, 17.6) | 4,022 (2,252, 5,791) | 245 | 15.1 (10.3, 21.6) |

Source: Kenya DHS 2022. Survey-weighted estimates; amounts are Kenyan shillings. The two n columns are the unweighted denominators for the columns to their right. Restricted to respondents who reported paying, which is the questionnaire's own skip pattern for these items: no amount or payer is recorded for a visit at which nothing was paid. Total cost is sh304; cash is sh305a, the part met out of pocket at the visit. The mean cash figure retains respondents who paid nothing in cash and the preceding column gives their share. Insurer contribution is any non-zero amount met by NHIF or private insurance. KDHS does not collect household consumption, so catastrophic health expenditure cannot be constructed from these data.

**Table 6.** Survey-weighted adjusted prevalence ratios for uninsured status and for any payment at the last outpatient visit within the population of adults meeting the Washington Group Short Set disability threshold, KDHS 2022.

| Characteristic | Uninsured APR (95% CI); p | Any payment APR (95% CI); p |
| --- | --- | --- |
| Insurance status |  |  |
| Any insurance (ref: uninsured) |  | 0.98 (0.92, 1.04); p = 0.532 |
| Sex |  |  |
| Women (ref: men) | 1.00 (0.96, 1.05); p = 0.876 | 1.00 (0.96, 1.05); p = 0.908 |
| Age group (ref: 18-29) |  |  |
| 30-44 | 0.89 (0.81, 0.97); p = 0.011 | 1.05 (0.95, 1.16); p = 0.331 |
| 45-59 | 0.75 (0.69, 0.81); p < 0.001 | 1.07 (0.97, 1.18); p = 0.177 |
| 60+ | 0.78 (0.73, 0.83); p < 0.001 | 1.05 (0.95, 1.16); p = 0.308 |
| Wealth quintile (ref: richest) |  |  |
| Richer | 1.44 (1.09, 1.91); p = 0.011 | 0.93 (0.83, 1.04); p = 0.180 |
| Middle | 1.85 (1.43, 2.41); p < 0.001 | 0.95 (0.86, 1.04); p = 0.283 |
| Poorer | 2.20 (1.70, 2.85); p < 0.001 | 0.97 (0.88, 1.07); p = 0.601 |
| Poorest | 2.35 (1.81, 3.05); p < 0.001 | 0.92 (0.82, 1.03); p = 0.127 |
| Place of residence |  |  |
| Rural (ref: urban) | 0.88 (0.81, 0.97); p = 0.007 | 1.03 (0.94, 1.12); p = 0.533 |
| Educational attainment (ref: no education) |  |  |
| Primary | 0.92 (0.87, 0.97); p = 0.002 | 1.02 (0.96, 1.09); p = 0.469 |
| Secondary | 0.82 (0.74, 0.91); p < 0.001 | 1.03 (0.95, 1.11); p = 0.479 |
| Higher | 0.67 (0.50, 0.90); p = 0.007 | 1.09 (1.00, 1.18); p = 0.052 |
| Disability severity |  |  |
| Severe functional difficulty (ref: moderate) | 1.02 (0.94, 1.11); p = 0.592 | 1.02 (0.95, 1.10); p = 0.563 |

Source: Kenya DHS 2022. Survey-weighted quasi-Poisson models with a log link; confidence limits use the survey degrees of freedom.
These models describe variation within the disability subpopulation; the disability contrast itself is in Table 3.
The payment model conditions on having used outpatient care, which is itself associated with disability, insurance and wealth, so its estimates are descriptive associations among users.

The cash payment separates into two parts that point in opposite directions. Insured respondents were more likely to pay nothing at all in cash: 11.5% (95% CI 7.3% to 17.6%) of insured payers with disability reported a zero cash amount, against 1.2% (95% CI 0.6% to 2.6%) of uninsured payers with disability, and in the adjusted model insurance was associated with a lower probability of any cash payment (PR 0.89, 95% CI 0.87 to 0.91; n = 7,031). Among those who did pay cash, insured respondents paid more (ratio of geometric means 1.20, 95% CI 1.06 to 1.34; n = 6,628), as did adults with disability (1.38, 95% CI 1.20 to 1.58). Retaining the zeros, mean cash paid was 4,022 KSh among insured payers with disability and 2,335 KSh among uninsured payers with disability (Table S5).

Among insured outpatient users with disability who reported paying and had a recorded payer split (n = 245), an insurer met part of the reported amount for 15.1% (95% CI 10.3% to 21.6%); the corresponding figure was 18.0% (95% CI 15.7% to 20.4%) and 18.0% (95% CI 15.7% to 20.4%) among insured adults without disability, and 17.7% (95% CI 15.6% to 20.0%) across all insured payers with payer data. These are survey-weighted proportions; the unweighted counts behind them are in Table S4. The mean cash share of the reported amounts among insured payers with disability was 86.2% (95% CI 81.1% to 91.4%). No uninsured respondent reported an insurer contribution.

These proportions are conditional on a payment having been reported. The payer items are not asked when nothing was paid, so contacts at which an insurer met the whole bill and contacts that were free at the point of use are indistinguishable and unobserved. Among insured outpatient users with disability, 34 of 324 reported no payment and 45 had no payer split. Assigning all of those contacts to “no insurer contribution” and then to “insurer contribution” bounds the proportion of insured contacts with any insurer contribution between 11.9% and 32.9% (Table S4). Both endpoints are survey-weighted estimates rather than known constants; their design-based 95% limits are 8.1% to 17.1% for the lower endpoint and 26.8% to 39.7% for the upper endpoint, which describe sampling uncertainty in each endpoint separately and are not a confidence interval for the partially identified quantity. Nothing between those bounds is identified by these data.

### 3.6 Variation within the disability population

Within the population of adults at the WG threshold (2,813), the largest contrasts in the adjusted model for uninsurance were across wealth quintiles: compared with the richest quintile, uninsurance was higher in the richer (APR 1.44 (95% CI 1.09 to 1.91)), middle (1.85 (95% CI 1.43 to 2.41)), poorer (2.20 (95% CI 1.70 to 2.85)) and poorest (2.35 (95% CI 1.81 to 3.05)) quintiles. Older age was associated with lower uninsurance, and severe difficulty was not associated with higher uninsurance than moderate difficulty (1.02 (95% CI 0.94 to 1.11); p = 0.592). In the corresponding payment model among outpatient users with disability (1,137), insurance was not associated with payment incidence (0.98 (95% CI 0.92 to 1.04); p = 0.532). These are contrasts within one subpopulation on a prevalence-ratio scale and are not comparable measures of importance across variables.

### 3.7 Sensitivity analyses

Grouping “don’t know” insurance responses with the uninsured left both the contrast adjusted for sex, age group and residence (PR 1.10, 95% CI 1.06 to 1.13) and the fully adjusted contrast (PR 1.01, 95% CI 0.98 to 1.03) unchanged. The domain-count specification of disability burden gave the same null as the severity summary after full adjustment. Excluding the internally inconsistent amount records did not change the amount estimates. Replicate-weight limits for the standardised differences agreed with the delta-method limits (Table S6). Adults excluded from the fully adjusted model for missing covariates differed little from those included.

## 4. Discussion

This analysis set out to describe, in one nationally representative adult sample, how insurance coverage, service use and payment at the point of care differ between adults who meet the Washington Group disability threshold and adults who do not, in the year before Kenya began replacing NHIF with the Social Health Authority. Five findings emerge, and the differences between them carry more weight than any one alone. Uninsurance was higher among adults at the threshold, by about 7 percentage points after adjustment for sex, age group and residence, and was attenuated to the null once wealth and education entered the model. The severity pattern was not a smooth gradient, and no step between adjacent categories survived full adjustment. Service use was higher at the threshold in every specification, for both outpatient contact and hospitalisation. Payment at the point of care was near-universal among outpatient users whether or not they held insurance coverage. And what an insurer contributed to that payment is only partly observable, because the survey does not ask who paid when no payment was reported.

The coverage finding is best read as a statement about how disability and economic position travel together rather than as an independent effect of functional difficulty. The attenuation is not produced by age, since the estimate adjusted for sex, age group and residence is essentially the unadjusted one; it appears only when wealth and education enter. Whether those two variables act here as prior confounders, as consequences of functional difficulty, or as both depends on when difficulty began, which KDHS does not record. The distinction is not merely technical. A systematic review of the relationship between disability and poverty in low- and middle-income countries found the association well supported but its direction rarely identified, for the same reason that constrains us: most of the evidence is cross-sectional and few surveys record onset [17]. Work on the direct costs of disability points the same way, finding extra costs that are sizeable and that rise with severity, which would place households containing an adult with functional difficulty lower in a wealth ranking partly because of the difficulty itself [18]. What can be said from these data is narrower than either reading: the coverage gap and household economic position are not separable here, and an analysis that adjusts for wealth and education is not thereby isolating the part of the gap that policy could address. That the socioeconomic patterning of coverage is pro-rich reproduces what earlier Kenyan and cross-country analyses have established [2–4]. The contribution of this study is the disability stratification and the joint treatment of coverage, use and payment in one adult sample, not the discovery that coverage follows wealth.

The severity result deserves separate emphasis because it corrects a reading these data might otherwise invite. Coverage does fall as severity rises, but the steps that reach significance depend on the adjustment set, none survives adjustment for wealth and education, and the domain-count specification gives the same answer. There is therefore no support here for a severity threshold at which the financing system fails, and the apparent gradient should not be read from the intervals alone. This matters for measurement as much as for policy: the WG-SS severity summary is a maximum across six domains rather than a validated scale, and treating it as an ordered exposure asks more of it than it was designed to carry.

Higher service use among adults at the threshold is consistent with greater need, although the survey measures neither morbidity nor unmet need, so need here is inferred rather than observed. Two features of the measure shape how the levels should be read. The four-week recall and the broad definition of an outpatient contact, which includes consultations at pharmacies and dispensaries, place these prevalences above what a narrower definition of a facility visit would give; the disability contrast rather than the level is the quantity of interest. The direction is nonetheless consistent with the international literature. A systematic review of access to general healthcare services for people with disabilities in low- and middle-income countries found utilisation frequently equal to or higher than among people without disabilities, alongside worse experiences of care and higher expenditure, and concluded that utilisation alone is a poor indicator of whether a health system is serving this population [19]. The WHO global report on health equity for persons with disabilities makes the same argument at greater scale, documenting persistent inequities in outcomes that coexist with substantial contact with services [20]. Our results fit that pattern: contact is not the binding constraint, and what happens at the contact is where the evidence here becomes uncomfortable.

The payment results should be read alongside the coarsened exact matching analysis of the same survey, which found higher gross expenditure among insured people and lower net out-of-pocket payment once reimbursement was taken into account [6]. Our gross finding agrees with theirs. Our cash findings agree in part, and only once the two parts are separated. Insured respondents were less likely to pay anything in cash, which points the same way as their net result, while conditional on paying cash the insured paid more, which does not. The two analyses use different samples and different estimands, ours conditional on a payment having been reported and theirs matched across those who paid nothing as well, so the comparison is limited. Both indicate that an insurance indicator and a payment indicator measure different things, and that a single coefficient on insurance will average over two opposing patterns. The lower probability of a positive cash payment among the insured is consistent with financial protection operating at the extensive margin, but it is not interpretable as an effect of insurance: coverage is not randomised, the analysis conditions on having sought care and on having reported a payment, and both conditions can induce selection. The larger amounts among insured people who did pay cash are as consistent with insured users reaching higher-level or costlier providers as with cover failing to protect them, and these data cannot separate the two.

That payment was near-universal is itself a finding, and it also limits what the payment models can show. Payment was reported by the great majority of outpatient users regardless of cover, so the payment-incidence outcome has little room to discriminate and its null should not be read as evidence that cover makes no difference to what households pay. The more informative quantity is what an insurer actually contributed, and that is precisely where KDHS stops observing: the amount and payer items are skipped whenever no payment was reported, so contacts at which an insurer met the whole bill are indistinguishable from contacts that were free at the point of use, and both are unobserved. Bounding rather than estimating that proportion is the honest response, and the bounds are wide. This is not a peculiarity of one survey. A recent systematic review of health insurance access among people with disabilities in low- and middle-income countries found a positive association with disability-specific service use but judged the evidence on general healthcare access and on financial protection inconclusive, in part because the available measures rarely capture what cover contributed at the point of care [21]. That review also reported coverage among people with disabilities averaging around two-fifths across the studies it assembled, which places the Kenyan figures reported here at the lower end of that range, though differences in disability measurement and in what counts as coverage make any such comparison indicative only.

Within Kenya, the qualitative finding that pro-poor financing schemes can enrol poor women with disabilities and still leave them paying at the facility [7] is consistent with what we observe at national scale, and it supplies a mechanism our data cannot: enrolment is not the same as benefit realisation. That distinction runs through the Kenyan record. NHIF’s own reform history documents repeated difficulty in converting membership into access [22], analyses of its purchasing arrangements identify weaknesses in provider payment and in the enforcement of benefit entitlements [23], evaluations among households with chronic conditions find members still exposed to substantial out-of-pocket costs [24], and facility-level work shows that the supply side is often not ready to deliver what a benefit package promises [25]. A second KDHS 2022 analysis reports coverage determinants in a 15-to-54 sample [5], but it draws on the individual and men’s recodes, in which the standard insurance variables are empty in this round, and it does not name the variable it used; we therefore do not treat its estimates as a benchmark for ours.

These observations bear on the SHA transition, though none is a prediction of what the reform will do and none is an evaluation of it: the data predate the transition entirely. Their value is as a pre-transition benchmark, against which SHA-era measurements of coverage, healthcare use and point-of-care payment can be compared. The Social Health Insurance Act 2023 and its regulations route support for people who cannot pay through means testing of the household rather than through a disability category [8,9]. The coverage deficit associated with disability is not separable from household economic position in these data, which is the dimension a means-tested premium subsidy targets, so the instrument is at least pointed at the right variable. At the same time, the WG-SS threshold used to identify disability in population surveys is not the instrument used to identify eligibility administratively, and KDHS records neither certification under the Persons with Disabilities Act nor programme enrolment [11]; nothing here indicates how many adults with functional difficulty a means test would in fact identify. On payment the data support a monitoring recommendation rather than a forecast. Enrolment status alone did not distinguish those who reported a payment from those who did not, and the payer information needed to say what cover contributed is missing for exactly the contacts where it would matter most. A monitoring framework that counts enrolments would therefore not detect a change in what households pay at the point of care, in either direction. The amount paid and the payer split are already collected in KDHS and would serve as indicators alongside utilisation, unmet need, benefit coverage and, in surveys that can measure it, catastrophic and impoverishing expenditure.

One further gap should be stated plainly rather than left to inference. KDHS 2022 does not record rehabilitation contacts, assistive product access or therapy costs, and this study makes no claim about them. That silence is consequential, because rehabilitation and assistive technology are where disability-specific need concentrates and where access in low- and middle-income countries is weakest [20,26]. Establishing whether access to those services moves with the SHA benefit package will require an instrument that asks about them, and no analysis of the current KDHS content, however careful, can substitute for one.

The strengths of this analysis are the national sampling frame, design-based estimation in which every subgroup is treated as a domain of one parent design rather than as a design rebuilt on filtered rows, the use of the WG-SS as the organising equity dimension, and the observation of coverage, use, amount and payer in one consistent adult sample. Partial identification is used where the data do not identify a point estimate, and the variance estimates for the standardised contrasts are checked against replicate weights constructed from the parent design.

The limitations are substantial and shape what can be concluded. The design is cross-sectional, with exposure and outcome measured at the same moment, so no causal claim is made and none of the reported associations should be read as an effect. Much of the information was reported by a household respondent rather than by the individual, and the person recode does not identify the informant for any record; proxy reporting is likely to matter most for the cognition and communication domains, for less visible difficulty, for healthcare use and payment, and for another member’s insurance status, and the direction of any resulting misclassification cannot be established from these data. The modules were also administered only in the long-questionnaire half of the sample. The available recode documentation does not establish a separate calibration adjustment for that split, so no weighted population totals are constructed; the two halves are closely similar on the five characteristics recorded in both, with a largest weighted difference of about one percentage point (Table S7), but agreement there does not establish agreement on the module variables themselves, which are unobserved in the short half, and residual selection bias cannot be excluded. No age at onset of functional difficulty is recorded, so the roles of education and wealth cannot be fixed and the attenuation in the third specification admits more than one explanation. Two analyses condition on care-seeking or on having paid, which can induce selection bias, and neither is interpretable as an effect of insurance. Most consequentially for the payment results, the amount and payer items are skipped when no payment was reported, so the payer composition of about a quarter of insured contacts is unobserved, and the insurer-contribution figure is bounded rather than estimated. KDHS collects no consumption or income measure, so catastrophic and impoverishing expenditure cannot be constructed, which rules out the financial-protection indicators this literature treats as standard. Severity is a maximum across domains rather than a validated scale, although the domain-count alternative gave the same answer. The seeing and hearing domains are further affected by the absence of usable information on whether respondents were assessed while using their usual assistive devices, since the glasses and hearing-aid variables are empty in this round; functional difficulty in those two domains may therefore be misclassified. Some cells are small, notably insured adults with severe difficulty (n = 19), and are marked rather than interpreted. Analyses are complete-case, though missingness was under 1%. The modules were administered in half of sampled households, so estimates generalise to adult usual residents represented by that half-sample design, and not to institutionalised populations, people absent from households, or those whose disability prevented reliable proxy report. Finally, the WG-SS threshold is not Kenyan administrative disability certification, and the two should not be treated as interchangeable.

## 5. Conclusions

Adults meeting the WG-SS disability threshold had higher healthcare utilisation and higher crude uninsurance than other adults in the KDHS 2022 long-questionnaire sample. The uninsurance difference was substantially attenuated after adjustment for wealth and education, whereas the differences in outpatient use and hospitalisation persisted under every adjustment set. Most outpatient users reported making a payment at their last visit regardless of insurance status. Because payer information was not collected when no payment was reported, the contribution of insurers could not be fully estimated and was bounded between 11.9% and 32.9% instead. These findings provide a pre-SHA benchmark showing why monitoring insurance enrolment alone may be insufficient: subsequent assessment should also examine healthcare use and payment at the point of care.

## Supporting information

This study is reported in accordance with the STROBE (Additional file 1) statement for cross-sectional studies.

The age threshold of 18 was set a priori to match the adult policy population for premium contribution, rather than the 15-year threshold used in the

## Data Availability

The Kenya Demographic and Health Survey 2022 microdata used in this study are available to registered researchers from The DHS Program following submission and approval of a data-access request. The data cannot be redistributed by the authors. Study-specific analysis scripts and supporting files are publicly available at https://github.com/gondamol/Kenya_DHS_Studies/tree/main/ST02_Disability_Insurance_Equity.

## List of abbreviations

APR: adjusted prevalence ratio
CI: confidence interval
DHS: Demographic and Health Survey
KDHS: Kenya Demographic and Health Survey
NHIF: National Hospital Insurance Fund
PR: person recode
SHA: Social Health Authority
WG-SS: Washington Group Short Set

## Additional files

Additional file 1: STROBE checklist.

Additional file 2: Supplementary tables. Table S1, participant flow and analysis denominators; Table S2, missing data; Table S3, prespecified contrasts between adjacent Washington Group severity categories; Table S4, bounds on insurer contribution; Table S5, two-part analysis of the cash payment; Table S6, delta-method and replicate-weight variance estimation; Table S7, long-against short-questionnaire adults on the characteristics recorded in both halves.

## Declarations

### Ethics approval and consent to participate

This study used de-identified secondary data from the Kenya Demographic and Health Survey 2022, obtained from The DHS Program under an approved data request. The original survey received ethical approval from the Kenya Medical Research Institute Scientific and Ethics Review Unit and the ICF Institutional Review Board. No new participant contact occurred in this secondary analysis and no additional ethical approval was required.

### Consent for publication

Not applicable.

### Availability of data and materials

KDHS 2022 microdata are available to registered researchers from The DHS Program following an approved data request at https://www.dhsprogram.com; they are not distributed by the authors. Study-specific analysis scripts and supporting files for this study are publicly available at https://github.com/gondamol/Kenya_DHS_Studies/tree/main/ST02_Disability_Insurance_Equity.

### Authors’ contributions

NWA conceptualised the study, conducted the statistical analysis, and drafted the manuscript. JO contributed to study design, interpretation of findings, and critical revision of the manuscript. Both authors approved the final version.

## Acknowledgements

The authors thank The DHS Program for access to KDHS 2022 microdata and the Kenya National Bureau of Statistics and the Ministry of Health for conducting the survey.

