## Supplementary material for "Insurance coverage, healthcare use, and outpatient payment among adults with disability in Kenya before the SHA transition: a secondary analysis of KDHS 2022": This study is reported in accordance with the STROBE (Additional file 1) statement for cross-sectional studies.

### STROBE checklist

Study type: Cross-sectional secondary analysis of KDHS 2022 person recode data.

Section names refer to the revised manuscript. Where an item is only partly addressable with these data, the entry says so rather than claiming coverage.

| Item | Reporting element | Location in manuscript |
| --- | --- | --- |
| 1 | Study design identified in title and abstract | Title; Abstract (Methods) |
| 2 | Background and rationale | Background |
| 3 | Objectives, stated as descriptive estimands | Background, final paragraph: three numbered objectives, with an explicit statement that no causal effect is estimated |
| 4 | Key elements of study design | Methods: Study design and data source, including the half-sample administration of the modules |
| 5 | Setting, survey context, and dates | Methods: Study design and data source |
| 6 | Eligibility criteria and participant selection | Methods: Study population; Table S1 |
| 7 | Definitions of outcomes, exposures and covariates | Methods: Variables (Disability; Insurance, service use and payment; Covariates and their assumed roles) |
| 8 | Data sources and measurement, including who reported | Methods: Study design and data source (household and largely proxy report); Methods: Variables |
| 9 | Potential sources of bias | Methods: Variables (the payer items are skipped when no payment was reported) and Statistical analysis (conditioning on care-seeking and on having paid); Discussion: Strengths and limitations (proxy report, selection, unobserved payer source, adjustment-set interpretation, complete-case analysis) |
| 10 | Study size | Methods: Study population; Results: Participant flow and sample characteristics; Table S1 |
| 11 | Handling of quantitative variables | Methods: Variables (severity categories, domain count, non-response codes for amounts); Methods: Statistical analysis (log transformation of amounts) |
| 12 | Statistical methods, confounding, subgroups, missing data, sensitivity analyses | Methods: Statistical analysis (one parent design with subgroup estimates taken as domains of it, adjustment sets, marginal standardisation with delta-method and replicate-weight limits, prespecified severity contrast family without multiplicity adjustment, two-part model for the cash payment, complete-case rule, four prespecified sensitivity analyses) |
| 13 | Participant flow | Results: Participant flow and sample characteristics; Table S1; comparison of included and excluded adults |
| 14 | Descriptive characteristics and missing data by variable | Results: Participant flow and sample characteristics; Table 1; Table S2 |
| 15 | Outcome data with denominators | Results, Sections 3.2 to 3.5; Tables 2, 4 and 5, each with outcome-specific denominators |
| 16 | Main results, unadjusted and adjusted, with precision | Results: The disability contrast under three adjustment sets; Table 3 (prevalence ratios and standardised differences); Table 6 (within-disability models); Figure 1 |
| 17 | Other analyses, including subgroups and sensitivity analyses | Results: Sensitivity analyses; Table S3 (prespecified severity contrasts); Table S4 (bounds on insurer contribution); Table S5 (two-part cash model); Table S6 (variance-estimator comparison); domain-count specification; don’t-know coding; exclusion of internally inconsistent amount records |
| 18 | Key results in relation to objectives | Discussion: Principal findings |
| 19 | Limitations, with direction of potential bias | Discussion: Strengths and limitations |
| 20 | Interpretation in the light of limitations and comparable evidence | Discussion: Relation to existing evidence; Implications for the SHA transition |
| 21 | Generalisability | Discussion: Strengths and limitations (half-sample design; household populations only) |
| 22 | Funding and role of funders | Front matter; Declarations: Funding |

#### Quantities that are bounded rather than estimated

The proportion of insured outpatient contacts at which an insurer met part of the cost is not identified. The amount and payer items are asked only of respondents who reported paying, so a contact at which an insurer met the whole bill and a contact that was free at the point of use are indistinguishable and unobserved. Table S4 reports the two extreme assumptions as bounds, and the manuscript does not report a point estimate for that quantity.

#### Items that these data cannot support

Catastrophic and impoverishing health expenditure are not reported. KDHS 2022 records the cost of a visit and the amounts met by each payer, but no household consumption or income measure, so no expenditure share can be constructed.

Disability certification status under the Persons with Disabilities Act, and enrolment in any premium subsidy programme, are not recorded in the person recode file. No statement is made about who would qualify for public support.

Rehabilitation and assistive-product access are not measured in KDHS 2022 and are not reported.
