## Supplementary material for "Insurance coverage, healthcare use, and outpatient payment among adults with disability in Kenya before the SHA transition: a secondary analysis of KDHS 2022": The age threshold of 18 was set a priori to match the adult policy population for premium contribution, rather than the 15-year threshold used in the

### Additional file 2: Supplementary tables

Table S1 traces the analytic sample from the full person recode file to the analytic base and then to each outcome-specific denominator, so that every count reported in the paper can be followed back to the file it came from. The three payment populations, which have different denominators, separate here for the first time.

Table S1. Participant flow from the KDHS 2022 person recode file to the analytic base and to each outcome-specific denominator.

| **Step** | **n** | **% of preceding population** |
| --- | --- | --- |
| Raw PR records | 156,571 |  |
| Adult usual residents aged 18+ with complete design variables | 77,909 | 49.8% |
| of whom in short-questionnaire households (modules not administered) | 37,625 | 48.3% |
| Adult usual residents in long-questionnaire households | 40,284 | 51.7% |
| Adults with a non-missing WG summary item (hdis9) | 40,240 | 99.9% |
| ST02 analytic base: adults with complete six-domain WG-SS data | 40,197 | 99.8% |
| with non-missing insurance status (sh27) | 39,911 | 99.3% |
| with non-missing outpatient use (sh31) | 40,063 | 99.7% |
| with non-missing hospitalisation (sh29) | 40,140 | 99.9% |
| outpatient users in the previous four weeks | 9,036 | 22.5% |
| outpatient users with a non-missing payment response (sh32) | 8,962 | 99.2% |
| outpatient users who paid, with a usable cost amount (sh304) | 7,267 | 81.1% |
| Payment analysis populations, which additionally require non-missing insurance status |  |  |
| outpatient users analysed | 8,908 | 22.2% |
| of whom payers with a usable cost amount | 7,228 | 81.1% |
| of whom payers with a recorded payer split | 7,027 | 97.2% |
| *Source: Kenya DHS 2022, person recode file. Unweighted counts.* | | |
| *Indented rows are subsets of the row above them. The analytic base is adults in long-questionnaire households with complete six-domain WG-SS data.* | | |

Table S2 gives the number and proportion of missing values for every outcome and adjustment variable within the analytic base. Missingness is under 1% on each variable, which is the basis for the complete-case approach used throughout.

Table S2. Missing values for each outcome and adjustment variable within the analytic base.

| **Variable** | **Denominator** | **Missing n** | **Missing %** |
| --- | --- | --- | --- |
| Insurance status (sh27) | 40,197 | 286 | 0.71 |
| Outpatient use (sh31) | 40,197 | 134 | 0.33 |
| Educational attainment | 40,197 | 114 | 0.28 |
| Hospitalisation (sh29) | 40,197 | 57 | 0.14 |
| Age group | 40,197 | 0 | 0.00 |
| Place of residence | 40,197 | 0 | 0.00 |
| Sex | 40,197 | 0 | 0.00 |
| WG severity | 40,197 | 0 | 0.00 |
| Wealth quintile | 40,197 | 0 | 0.00 |
| Payment at last outpatient visit (sh32), among outpatient users | 9,036 | 74 | 0.82 |
| *Source: Kenya DHS 2022. Unweighted counts within the analytic base.* | | | |
| *Analyses are complete-case within each model. Adults excluded from the fully adjusted insurance model for missing covariates are compared with those included in the Results.* | | | |

Table S3 reports the prespecified contrasts between adjacent severity categories for uninsured status under each adjustment set. It is the formal test behind the statement in the Results that the severity gradient depends on the model, and it is what the severity comparison should be read from rather than from the overlap of the intervals in Table 2.

Table S3. Prespecified contrasts between adjacent Washington Group severity categories for uninsured status.

| **Specification** | **Contrast** | **Prevalence ratio (95% CI); p** |
| --- | --- | --- |
| Crude | Mild vs no difficulty | 1.00 (0.98, 1.03); p = 0.722 |
| Crude | Moderate vs mild | 1.09 (1.05, 1.13); p < 0.001 |
| Crude | Severe vs moderate | 1.07 (0.98, 1.17); p = 0.110 |
| Crude | Severe vs no difficulty (extreme contrast) | 1.18 (1.08, 1.28); p < 0.001 |
| Adjusted for sex, age group, residence | Mild vs no difficulty | 1.03 (1.01, 1.06); p = 0.018 |
| Adjusted for sex, age group, residence | Moderate vs mild | 1.07 (1.03, 1.11); p < 0.001 |
| Adjusted for sex, age group, residence | Severe vs moderate | 1.05 (0.96, 1.14); p = 0.265 |
| Adjusted for sex, age group, residence | Severe vs no difficulty (extreme contrast) | 1.16 (1.07, 1.25); p < 0.001 |
| Additionally adjusted for wealth, education | Mild vs no difficulty | 1.00 (0.98, 1.02); p = 0.967 |
| Additionally adjusted for wealth, education | Moderate vs mild | 1.01 (0.98, 1.04); p = 0.645 |
| Additionally adjusted for wealth, education | Severe vs moderate | 1.02 (0.93, 1.11); p = 0.710 |
| Additionally adjusted for wealth, education | Severe vs no difficulty (extreme contrast) | 1.02 (0.94, 1.11); p = 0.579 |
| *Source: Kenya DHS 2022. Survey-weighted quasi-Poisson models with a log link; contrasts estimated on the linear predictor and exponentiated, with confidence limits on the survey degrees of freedom.* | | |
| *Design-based global Wald tests of the severity term are reported in the Results.* | | |

Table S4 gives the bounds on the proportion of insured outpatient contacts at which an insurer met any part of the cost, together with the counts the bounds rest on. Because the payer items are skipped whenever no payment was reported, the quantity is partially identified: the table reports the two assumptions that bound it and design-based limits for each endpoint, not a point estimate.

Table S4. Bounds on the proportion of insured outpatient contacts at which an insurer met any part of the cost.

| **Population** | **Outpatient users** | **Reported no payment** | **Payer split missing** | **Insurer contribution recorded** | **Bounds on insurer contribution** | **BoundsCI** |
| --- | --- | --- | --- | --- | --- | --- |
| All insured outpatient users | 3,014 | 381 | 402 | 441/2,231 (19.8%) | 13.7% to 36.0% | 12.1% to 15.6%; 33.7% to 38.3% |
| Insured outpatient users with disability | 324 | 34 | 45 | 38/245 (15.5%) | 11.9% to 32.9% | 8.1% to 17.1%; 26.8% to 39.7% |
| *Source: Kenya DHS 2022. Counts are unweighted; both bounds are survey-weighted proportions estimated on the parent design with the DHS person weights.* | | | | | | |
| *Lower bound: every contact at which nothing was paid, and every contact with a missing payer split, is treated as having no insurer contribution. Upper bound: all of them are treated as met by an insurer.* | | | | | | |
| *The payer items are not asked of respondents who reported no payment, so no estimate between these bounds is identified by these data.* | | | | | | |
| *The final column gives design-based 95% limits for the lower and upper endpoint separately. They describe sampling uncertainty in each endpoint and are not a confidence interval for the partially identified quantity.* | | | | | | |

Table S5 sets out the two-part analysis of the cash payment. Part one is the probability of paying anything in cash, which is where the zero-cash records sit, and part two is the amount among those who paid something. The two parts are reported together because either one alone describes a different quantity from the one the policy question asks about.

Table S5. Two-part analysis of the cash payment at the last outpatient visit, among adults who reported paying.

| **Part** | **Term** | **Estimate (95% CI); p** | **Scale** |
| --- | --- | --- | --- |
| Any cash paid (n = 7,032) | WG disability threshold | 0.99 (0.98, 1.01); p = 0.545 | Prevalence ratio |
| Any cash paid (n = 7,032) | Any insurance | 0.89 (0.87, 0.91); p < 0.001 | Prevalence ratio |
| Amount, given cash paid (n = 6,628) | WG disability threshold | 1.38 (1.20, 1.58); p < 0.001 | Ratio of geometric means |
| Amount, given cash paid (n = 6,628) | Any insurance | 1.20 (1.06, 1.34); p = 0.003 | Ratio of geometric means |
| *Source: Kenya DHS 2022. Survey-weighted models adjusted for sex, age group, residence, wealth quintile and educational attainment; confidence limits use the survey degrees of freedom.* | | | |
| *Part one is the probability of paying anything in cash, which is where insurance appears as a zero; part two is the amount among those who paid something in cash and is conditional on that.* | | | |

Table S6 places the delta-method and replicate-weight confidence limits for the standardised prevalence differences side by side. The point estimates are identical by construction and only the variance estimator differs, so the table shows how much the model-based limits depend on treating the weighted covariate distribution as fixed.

Table S6. Standardised prevalence differences under delta-method and bootstrap replicate-weight variance estimation.

| **Outcome** | **Specification** | **Delta-method difference (95% CI)** | **Replicate-weight difference (95% CI)** |
| --- | --- | --- | --- |
| Uninsured | Crude | +7.1 (+4.7, +9.5) | +7.1 (+4.7, +9.4) |
| Uninsured | Adjusted for sex, age group, residence | +6.7 (+4.4, +9.0) | +6.7 (+4.4, +9.0) |
| Uninsured | Additionally adjusted for wealth, education | +0.6 (-1.3, +2.5) | +0.6 (-1.3, +2.5) |
| Outpatient use | Crude | +20.1 (+17.7, +22.5) | +20.1 (+17.8, +22.4) |
| Outpatient use | Adjusted for sex, age group, residence | +12.5 (+10.2, +14.7) | +12.5 (+10.3, +14.6) |
| Outpatient use | Additionally adjusted for wealth, education | +14.0 (+11.7, +16.3) | +14.0 (+11.7, +16.2) |
| Hospitalisation | Crude | +8.3 (+6.6, +10.0) | +8.3 (+6.6, +10.0) |
| Hospitalisation | Adjusted for sex, age group, residence | +7.2 (+5.4, +9.0) | +7.2 (+5.4, +8.9) |
| Hospitalisation | Additionally adjusted for wealth, education | +7.7 (+5.9, +9.6) | +7.7 (+5.9, +9.5) |
| *Source: Kenya DHS 2022. Point estimates are identical by construction; only the variance estimator differs.* | | | |
| *Delta-method limits treat the weighted covariate distribution as fixed. Replicate-weight limits refit the model in 500 subbootstrap replicates constructed from the parent design, with the analytic domain applied after the replicate weights are formed.* | | | |

Table S7 compares long-questionnaire and short-questionnaire adults on the five characteristics recorded in both halves of the sample. Because the disability, insurance, utilisation and payment modules were administered only in long-questionnaire households, every estimate in this study is conditional on that half; this table is the check on how far that restriction moves the composition of the sample. It cannot speak to the module variables themselves, which are not observed in the short half.

Table S7. Survey-weighted composition of long-questionnaire and short-questionnaire adults on the characteristics recorded in both halves.

| **Characteristic** | **Long questionnaire, %** | **Short questionnaire, %** | **Difference, pp** | **p** |
| --- | --- | --- | --- | --- |
| Age group |  |  |  | 0.470 |
| 18-29 | 37.4 | 38.1 | -0.7 |  |
| 30-44 | 31.9 | 31.5 | +0.4 |  |
| 45-59 | 17.6 | 17.4 | +0.2 |  |
| 60+ | 13.2 | 13.0 | +0.2 |  |
| Education |  |  |  | 0.534 |
| No education | 9.4 | 9.1 | +0.4 |  |
| Primary | 36.8 | 36.8 | -0.0 |  |
| Secondary | 35.2 | 35.6 | -0.4 |  |
| Higher | 18.6 | 18.6 | +0.1 |  |
| Residence |  |  |  | 0.186 |
| Urban | 37.3 | 37.9 | -0.6 |  |
| Rural | 62.7 | 62.1 | +0.6 |  |
| Sex |  |  |  | 0.018 |
| Men | 47.2 | 48.1 | -0.8 |  |
| Women | 52.8 | 51.9 | +0.8 |  |
| Wealth quintile |  |  |  | 0.360 |
| Richest | 22.8 | 23.0 | -0.2 |  |
| Richer | 22.5 | 21.7 | +0.9 |  |
| Middle | 19.8 | 20.6 | -0.8 |  |
| Poorer | 18.5 | 18.4 | +0.0 |  |
| Poorest | 16.4 | 16.3 | +0.1 |  |
| *Source: Kenya DHS 2022. Survey-weighted column percentages estimated on the parent design.* | | | | |
| *P values are design-based Rao-Scott F tests of independence between questionnaire half and the characteristic, reported once per characteristic.* | | | | |
| *Agreement here does not establish that the two halves would agree on the module variables, which are unobserved in the short half.* | | | | |
